# Whole-genome sequencing reveals new Alzheimer’s disease-associated rare variants in loci related to synaptic function and neuronal development

**DOI:** 10.1101/2020.11.03.20225540

**Authors:** Dmitry Prokopenko, Sarah L. Morgan, Kristina Mullin, Oliver Hofmann, Brad Chapman, Rory Kirchner, Sandeep Amberkar, Inken Wohlers, Christoph Lange, Winston Hide, Lars Bertram, Rudolph E. Tanzi

## Abstract

**INTRODUCTION:** Genome-wide association studies have led to numerous genetic loci associated with Alzheimer’s disease (AD). Whole-genome sequencing (WGS) now permit genome-wide analyses to identify rare variants contributing to AD risk.

**METHODS:** We performed single-variant and spatial clustering-based testing on rare variants (minor allele frequency ≤1%) in a family-based WGS-based association study of 2,247 subjects from 605 multiplex AD families, followed by replication in 1,669 unrelated individuals.

**RESULTS:** We identified 13 new AD candidate loci that yielded consistent rare-variant signals in discovery and replication cohorts (4 from single-variant, 9 from spatial-clustering), implicating these genes: FNBP1L, SEL1L, LINC00298, PRKCH, C15ORF41, C2CD3, KIF2A, APC, LHX9, NALCN, CTNNA2, SYTL3, CLSTN2.

**DISCUSSION:** Downstream analyses of these novel loci highlight synaptic function, in contrast to common AD-associated variants, which implicate innate immunity. These loci have not been previously associated with AD, emphasizing the ability of WGS to identify AD-associated rare variants, particularly outside of coding regions.

## Introduction

Alzheimer’s disease (AD) is the most common neurodegenerative disorder and one of the most challenging societal problems in the industrialized world. Susceptibility to AD is determined by both monogenic and polygenic risk factors as well as environmental exposure. Monogenic AD most often presents as early-onset (<60 years) familial AD (EOFAD), constituting less than 5% of all cases, and caused by any of hundreds of very rare mutations in at least three genes: amyloid precursor protein (*APP*), presenilin 1 (*PSEN1*) and presenilin 2 (*PSEN2*). The vast majority of AD cases are sporadic or familial late-onset (>60 years) AD (LOAD) and are of a genetically complex, polygenic background with contributions from both genetic and non-genetic factors. The identification of genetic determinants underlying polygenic AD has been the aim of more than one thousand genetic association studies^1^, including more than 75 genome-wide association studies (GWAS) on AD and related traits as outcomes (according to EBI’s GWAS catalog^2^; https://www.ebi.ac.uk/gwas/). The largest AD GWAS^3^ to date was conducted on over 600,000 individuals and highlighted a total of 29 independent genome-wide significant (*P*<5×10-8) AD risk loci^4^, while the recent GWAS by Kunkle *et al*.^5^ found 25 loci in their analyses of clinically diagnosed LOAD in over 90,000 individuals. Essentially, these and other AD GWAS focused on common (typically with a minor allele frequency [MAF] ≥1%) variants either directly assayed or imputed using high-density reference panels. The few exceptions to these common-variant studies utilized either microarray-based or next-generation sequencing (NGS)-based genotyping limited to exonic variants and identified rare (MAF<1%) missense variants either increasing (*TREM2, PLCG2, ABI3, ADAM10*) or decreasing (*APP, CD33*) risk for AD^6–9^.

In this study, we performed deep (>40x) whole-genome sequencing (WGS) to search for novel AD variants in 2,247 individuals from 605 multiplex AD families from the National Institute on Mental Health (NIMH)^10^ and National Institute on Aging (NIA) Alzheimer’s Disease Sequencing Project (ADSP)^11^ data sets. Analyses were focused on rare variants with MAF <1% (based on the non-Finnish European subset of gnomAD v3^12^, unless stated otherwise) and entailed single-variant and spatial clustering-derived (i.e. “region-based”) testing. Suggestive findings (*P*<5×10-4) were validated in publicly available WGS (NIA ADSP case-control population^13^) data on more than 1,650 independent AD cases and controls. In total, we highlight 4 single-variant and 9 region-based findings exhibiting consistent rare-variant association with AD across the discovery and replication phases in our study. None of the newly implicated loci were previously highlighted in any of the common-variant AD GWAS. Functionally, our results extend existing knowledge on the underlying disease pathways highlighted by common variants and converge upon a role for neuroplasticity and synaptic function, emphasizing the power of WGS in the context of rare-variant-based gene discovery efforts.

## Results

### Description of general sequencing metrics

After sample quality control (QC) (see Methods), WGS data from 2,247 individuals (NIMH n=1,393 and NIA ADSP families n=854; hereafter referred to as the “discovery sample” (Supplementary Table 1; see Fig. 1 for an overview of the study design) was available for subsequent analyses. Median read depth across the genome in NIMH was 40.4-fold (mean 41.2). Within the discovery sample, we identified a total of 54,669,406 sequence variants, of which 40,542,616 were listed in the non-Finnish European subset of the Genome Aggregation Database (gnomAD [URL: gnomad.broadinstitute.org]; v3, n=32,399, Supplementary Fig. 1). 907,273 (2%) of these were located in protein-coding exons (Supplementary Fig. 2). Of all identified variants, the vast majority, i.e. 31,200,539 (77%) were “rare” (MAF ≤1%), while 2,855,054 (7%) were “infrequent” (≤5% MAF >1%), and 6,487,023 (16%) were “common” (MAF >5%). Overall, we captured a large proportion of the “common” (95.8%) and “infrequent” (90.9%) variant space, using gnomAD as reference. As expected, the captured proportion was smaller for “rare” variants (11.7%), which can be attributed to the difference in sample sizes. After variant QC (Methods), 18,263,694 variants, 11,012,452, of which were rare, were used in subsequent analyses.

**Table 1.**
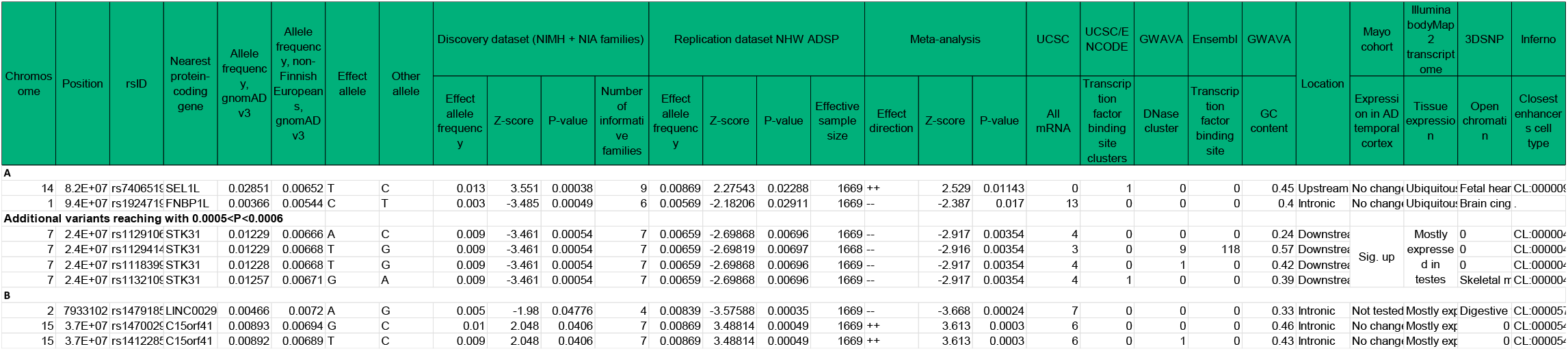
Top single-variant AD association results. **a**, Single-variant AD association results with *P*<0.0005 (*P*<0.0006) in the discovery dataset and consistent (i.e. *P*<0.05 and same direction of effect) association in ADSP NHW WGS replication dataset. **b**, Single-variant AD association results with consistent (i.e. *P*<0.05 and same direction of effect in discovery and replication dataset) association at *P*<0.0005 after meta-analysis.

**Fig. 1.**
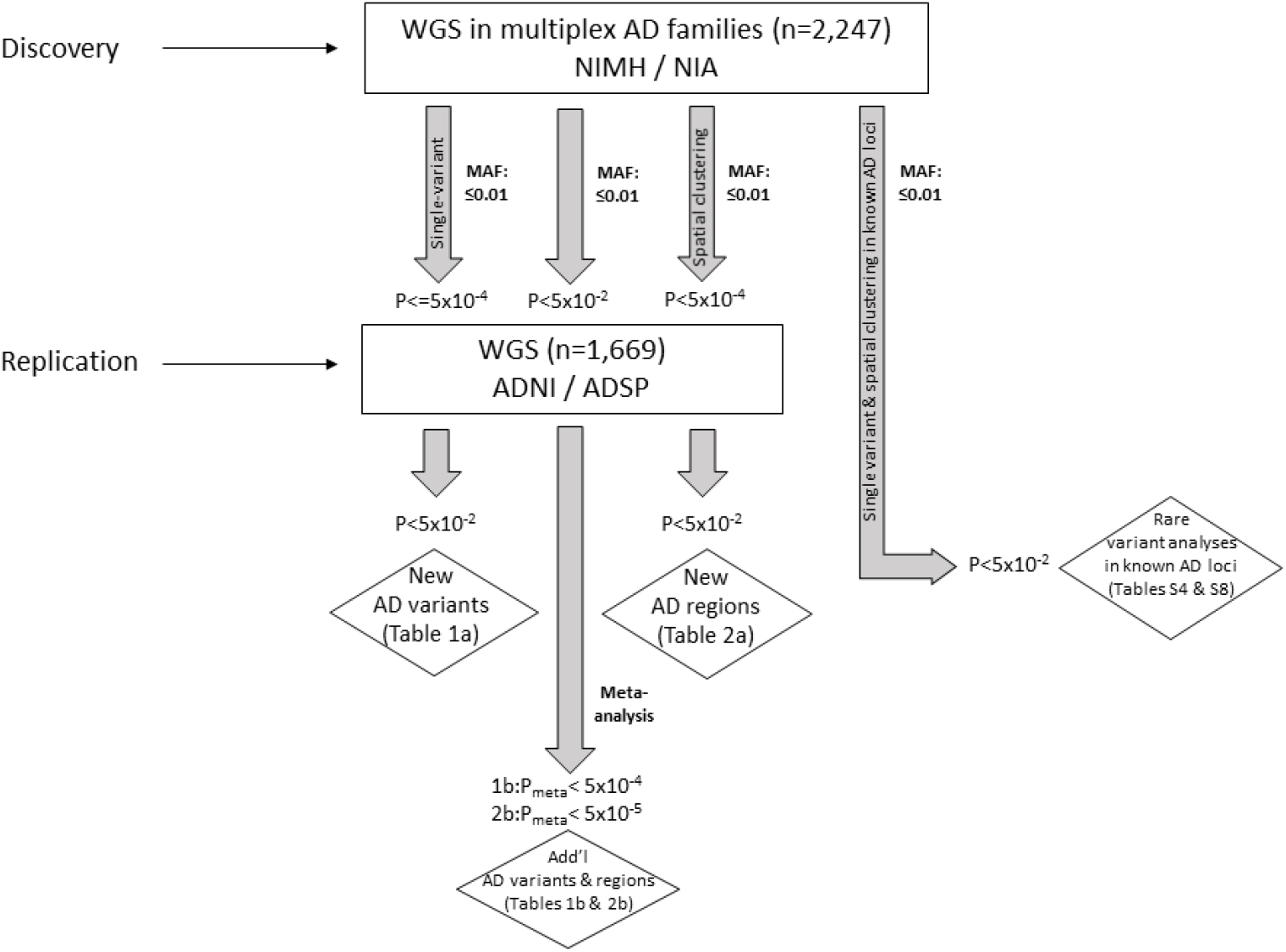
Data analysis workflow.

**Fig. 2.**
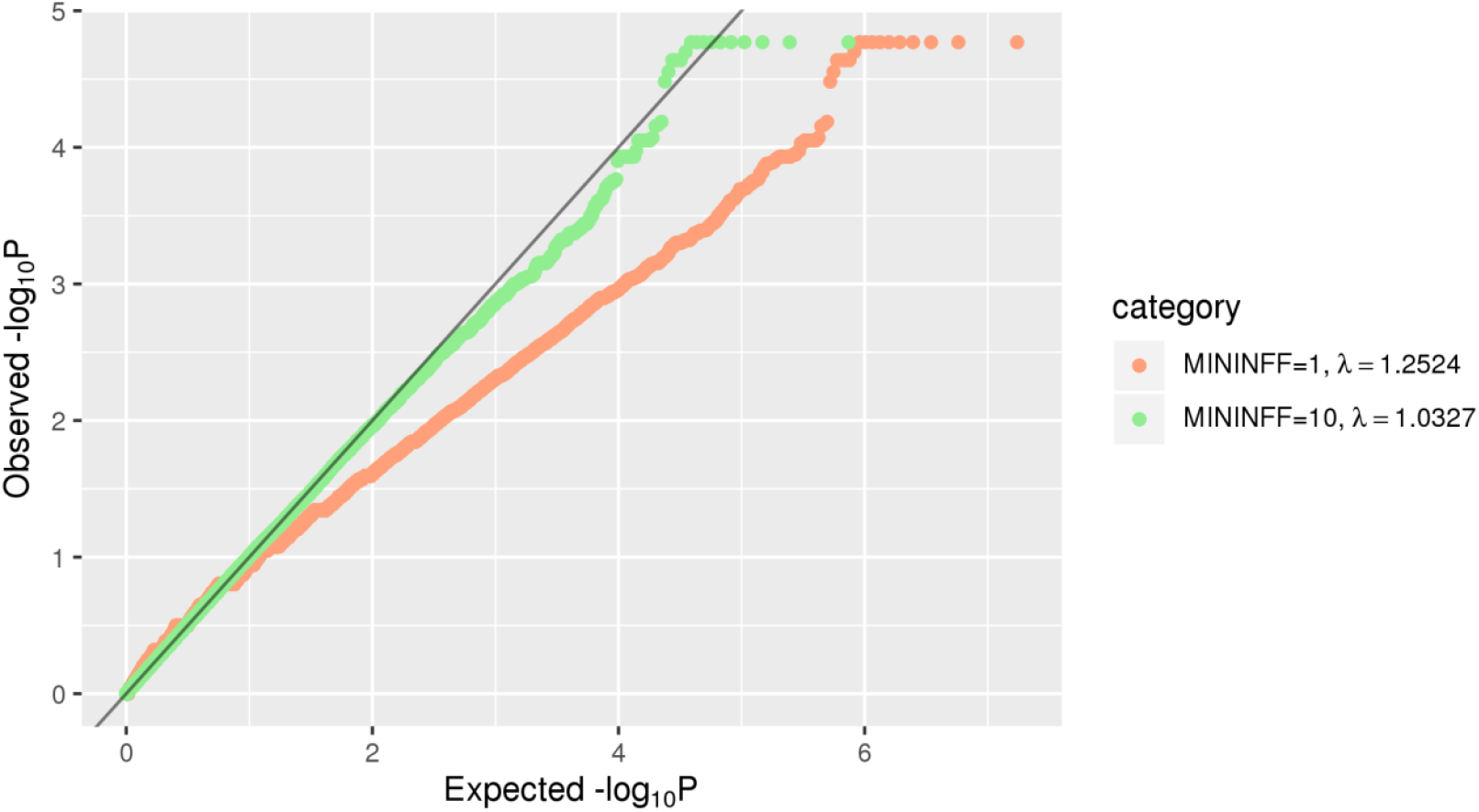
QQ plot of rare (MAF<=0.01) single-variant association results in the family-based discovery dataset (NIMH and NIA cohorts). Red line corresponds to all statistics, where at least one informative family is observed. Green line corresponds to statistics with at least ten informative families.

### Single-variant AD association results

To probe for association between single markers and AD status, we used the FBAT Toolkit^14^ in the family-based discovery dataset and logistic regression in the case-control replication data (Methods). These analyses revealed a total of 24,301 rare variants showing association with AD at *P*<0.01. As can be seen from the corresponding QQ plot (Fig. 2), we observed a deflation of test statistics starting from *P*<0.05. This deflation can be attributed to the fact that the FBAT-statistics is conservative in the case of a small number of informative families and/or low allele frequencies. Of the variants showing association at *P*<0.01, a total of 271 attained *P*<5×10-4 (Supplementary Table 2) and were prioritized for validation assessments in the independent WGS case-control dataset (NIA ADSP non-Hispanic whites (NHW), n=1669; hereafter referred to as “replication dataset”; Fig. 1). These assessments converged on two variants in two regions (rs74065194 approx. 200kb downstream from *SEL1L* [MAF = 0.0066; *P*_meta_ = 0.011] and rs192471919 intronic of *FNBP1L* [MAF = 0.0054; *P*_meta_ = 0.017]) to show at least nominal replication with the same direction of effect as in the discovery datasets (Fig. 3a, Table 1a). In addition, we highlight four variants which yielded *P* = 0.000538, i.e. just above our screening threshold, located approx. 100kb downstream of *STK31* [MAF = 0.0067; *P*_meta_ = 0.0035].

**Table 2.** Top spatial-clustering-based AD association results. **a**, Replicated spatial-clustering-based AD association results for regions showing *P*<0.0005 in the discovery dataset. **b**, Top spatial-clustering-based AD association results based on meta-analysis (P_meta_ < 5×10-5 and P_discovery_ < 0.05). The *PRKCH* region was identified in both arms of the regional study, hence appears in both: a and b. Overlapping/GREAT-assigned gene did not differ from the nearest gene assignment. *significantly more annotations than expected by chance after correcting for multiple testing.

| Chromosome | First SNV in the region (chr.pos:ref:alt) | Last SNV in the region (chr.pos:ref:alt) | Nearest protein-coding gene | Discovery dataset (NIMH + NIA families) |  | Replication dataset NHW ADSP |  | Meta-analysis | Location | Mayo | Illumina bodyMap 2 transcriptome | FANTOM | FANTOM | Ensembl | Ensembl | Ensembl | Ensembl | Ensembl | Ensembl | UCSC | UCSC | UCSC |
| --- | --- | --- | --- | --- | --- | --- | --- | --- | --- | --- | --- | --- | --- | --- | --- | --- | --- | --- | --- | --- | --- | --- |
|  |  |  |  | Number of SNVs in the region | P-value | Number of SNVs in the region | P-value | P-value |  |  |  |  |  |  |  |  |  |  |  |  |  |  |
| A |  |  |  |  |  |  |  |  |  |  |  |  |  |  |  |  |  |  |  |  |  |  |
| 14 | 14:611767 | 14:611880 | PRKCH | 53 | 2.51E-05 | 53 | 0.021069 | 8.17E-06 | Upstream | Sig. lower | Mostly exp | 86 | 6 | 14 | 10 | 0 | 61* | 0 | 9 | 81 | 0 | 17 |
| 11 | 11:740251 | 11:740313 | C2CD3 | 47 | 8.36E-05 | 42 | 0.045428 | 5.12E-05 | Intronic an | No change | Ubiquitous | 0 | 3 | 0 | 0 | 0 | 15 | 0 | 2 | 12 | 0 | 8 |
| 5 | 5:6197263 | 5:6197579 | KIF2A | 13 | 0.000169 | 16 | 0.046496 | 0.0001 | Upstream | No change | Mostly exp | 0 | 0 | 0 | 0 | 0 | 0 | 0 | 0 | 4 | 0 | 1 |
| 5 | 5:1128195 | 5:1128273 | APC | 25 | 0.000337 | 29 | 0.043747 | 0.000179 | Intronic an | No change | Mostly exp | 0 | 5 | 0 | 10 | 0 | 0 | 0 | 0 | 15 | 0 | 5 |
| B |  |  |  |  |  |  |  |  |  |  |  |  |  |  |  |  |  |  |  |  |  |  |
| 14 | 14:611767 | 14:611880 | PRKCH | 53 | 2.51E-05 | 53 | 0.021069 | 8.17E-06 | Upstream | Sig. lower | Mostly exp | 86 | 6 | 14 | 10 | 0 | 61* | 0 | 9 | 81 | 0 | 17 |
| 1 | 1:1979175 | 1:1979292 | LHX9 | 44 | 0.007622 | 39 | 0.000131 | 1.48E-05 | Intronic an | Not tested | Mostly exp | 429* | 24 | 1 | 0 | 0 | 104 | 15* | 31 | 117 | 2 | 15 |
| 13 | 13:101143 | 13:101164 | NALCN | 106 | 0.002585 | 134 | 0.000427 | 1.63E-05 | Intronic an | No change | Mostly exp | 0 | 2 | 2 | 26 | 1 | 0 | 19* | 73* | 31 | 0 | 18 |
| 2 | 2:7985374 | 2:7985689 | CTNNA2 | 15 | 1.22E-05 | 18 | 0.154028 | 2.67E-05 | Intronic | No change | Mostly exp | 0 | 0 | 1 | 6 | 0 | 0 | 0 | 19 | 7 | 0 | 3 |
| 6 | 6:1586776 | 6:1586834 | SYTL3 | 29 | 0.017664 | 28 | 0.000116 | 2.9E-05 | Intronic an | Not tested | Ubiquitous | 0 | 0 | 0 | 0 | 1 | 0 | 0 | 0 | 6 | 0 | 3 |
| 3 | 3:1403989 | 3:1404496 | CLSTN2 | 233 | 1.19E-05 | 273 | 0.196429 | 3.27E-05 | Intronic an | No change | Mostly exp | 0 | 9 | 7 | 32* | 2 | 57 | 0 | 16 | 40 | 0 | 44 |

**Fig. 3.**
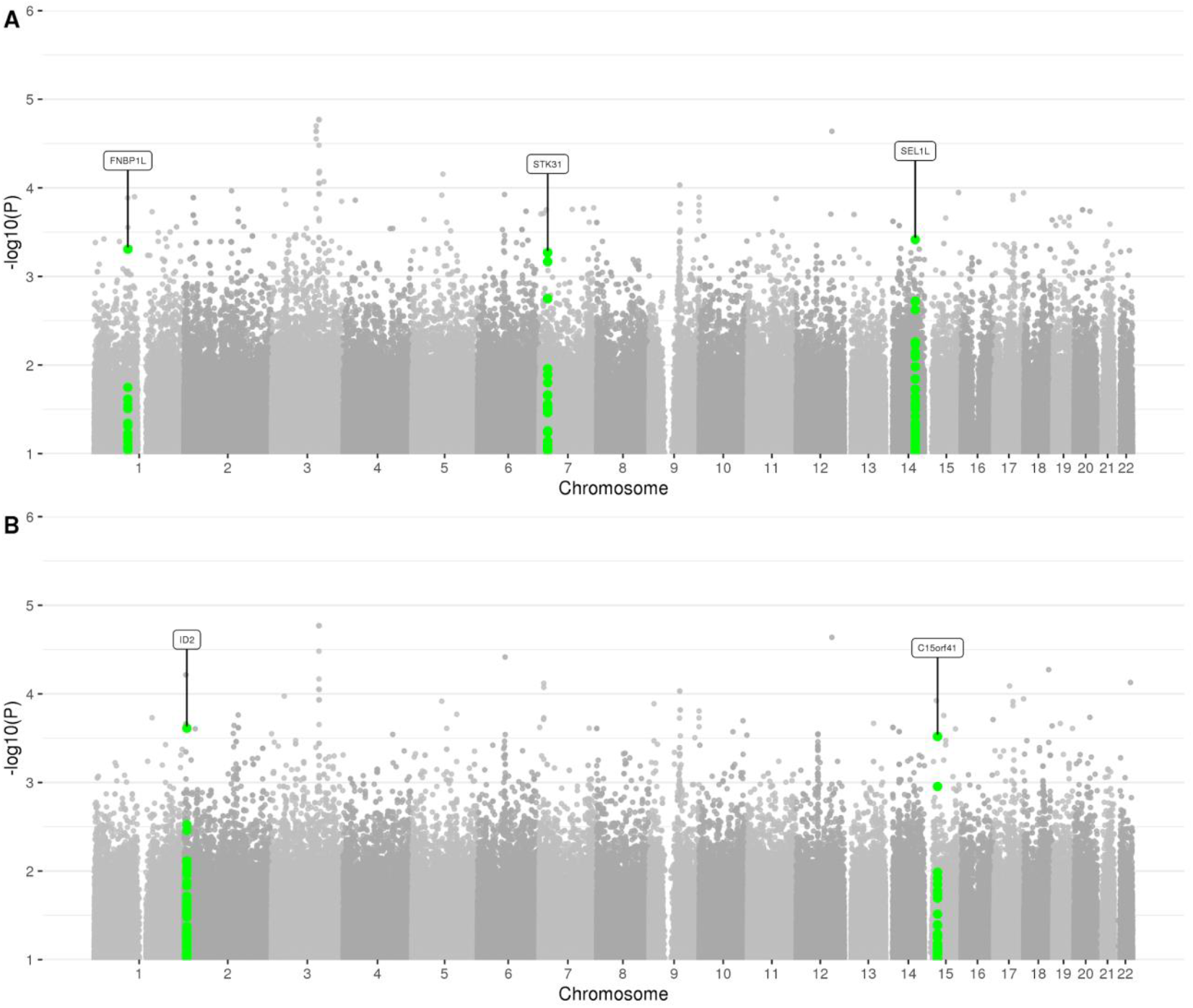
Manhattan plot of rare (MAF<=0.01) single-variant association results in the family-based discovery dataset (NIMH and NIA cohorts). Genes which correspond to replicated variants as described in the workflow (Fig. 1) are highlighted.

In a second filtering paradigm, we selected variants showing consistent (i.e. *P*_discovery_<0.05 *and* same direction of effect in discovery and replication datasets) association at *P*<0.0005 following meta-analysis. This revealed three additional single variant associations in two loci (i.e. rs147918541 intronic of *LINC00298* and approx. 700kb upstream of *ID2* [MAF = 0.0072; *P*_meta_ = 2.44×10-4], and rs147002962 and rs141228575, both intronic of *C15orf41* [MAF = 0.0069; *P*_meta_ = 3.03×10-4]; Fig. 3b; Table 1b). Furthermore, we assessed the recently described^9^ “exome-chip”-based rare-variant association signals in *TREM2* as well as *PLCG2* and *ABI3* (Supplementary Table 3). This revealed significant association with one of the two *TREM2* variants (rs75932628 [MAF = 0.0021; *P*_meta_ = 0.0329) as well as suggestive support for rs72824905 in *PLCG2* in the discovery sample only [MAF = 0.0087; *P*_discovery_ = 0.0546, *P*_meta_ = 0.259]). In contrast, we did not observe evidence for association with the second *TREM2* variant (rs143332484) or rs616338 in *ABI3* in either the discovery or the replication samples. Finally, we identified at least 786 nominally (*P*<0.05) significant rare-variant signals in genes corresponding to loci previously associated with AD in common-variant GWAS^3,5^ (Supplementary Table 4) suggesting that at least some of the common-variant signals in these loci can be attributed to rare sequence variation (in line with earlier findings^15,16^). For comparison, we also plotted single-variant association results in the discovery cohorts without MAF restriction, i.e. for both rare and common variants (Supplementary Fig. 3 and Supplementary Fig. 4) and compared these with the 29 GWAS SNPs from Jansen *et al*.^3^ (Supplementary Table 5) and 25 GWAS SNPs from Kunkle *et al*.^5^ (Supplementary Table 6). As expected, these analyses revealed a pronounced, genome-wide significant (P<5×10-8) signal with markers in the *APOE* region on chromosome 19q13 as well as suggestive signals with several of the other common-variant GWAS signals.

**Fig. 4.**
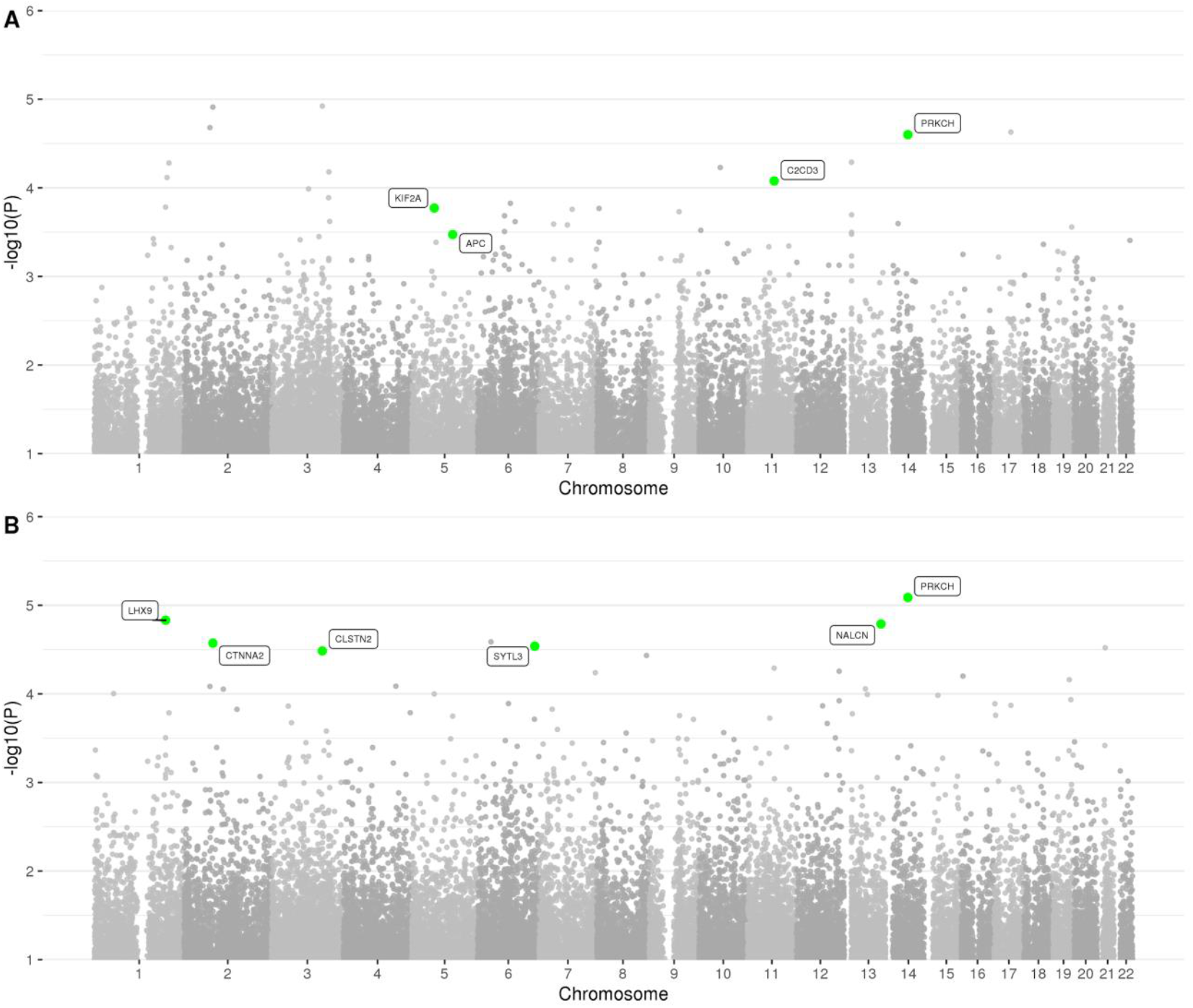
Manhattan plot of spatial clustering association results based on rare (MAF<=0.01) variants in the family-based discovery dataset (NIMH and NIA cohorts). Highlighted are genes, which correspond to replicated regions, described in the workflow (Fig. 1).

### Spatial-clustering AD-association results

Our second analysis arm computed aggregated results on consecutive runs of rare variants in the discovery dataset. In principle, this is similar to “gene-based” testing (such as performed by VEGAS^17^ or MAGMA^18^) except the approach applied here^19^ utilizes *all* available variants, including those located between genes that are otherwise typically omitted from this type of analysis, e.g. VEGAS. These analyses revealed a total of 1,756 regions showing association with AD at *P*<0.01 (for a Manhattan and QQ plot of all spatial-clustering-based rare-variant results see Fig. 4 A and B and Fig. 5). Using *P*<5×10-4 as threshold yielded signals in 47 regions in the discovery datasets (Supplementary Table 7), four of which also showed at least nominal evidence for independent replication in the NHW ADSP dataset (*PRKCH* [*P*_meta_ = 8.17×10-6], *C2CD3* [*P*_meta_ = 5.12×10-5], *KIF2A* [*P*_meta_ = 1.00×10-4], *APC* [*P*_meta_ = 1.79×10-4]; Table 2a). A further six (five of which were novel) candidate gene regions (*PRKCH, LHX9, NALCN, CTNNA2, SYTL3, CLSTN*) were highlighted in the secondary analyses focusing on top meta-analysis results (*P*_meta_ < 5×10-5 and *P*_discovery_ < 0.05) only, yielding association signals with *P*-values ranging from 3.27×10-5 to 8.17×10-6 (Table 2b).

**Fig. 5.**
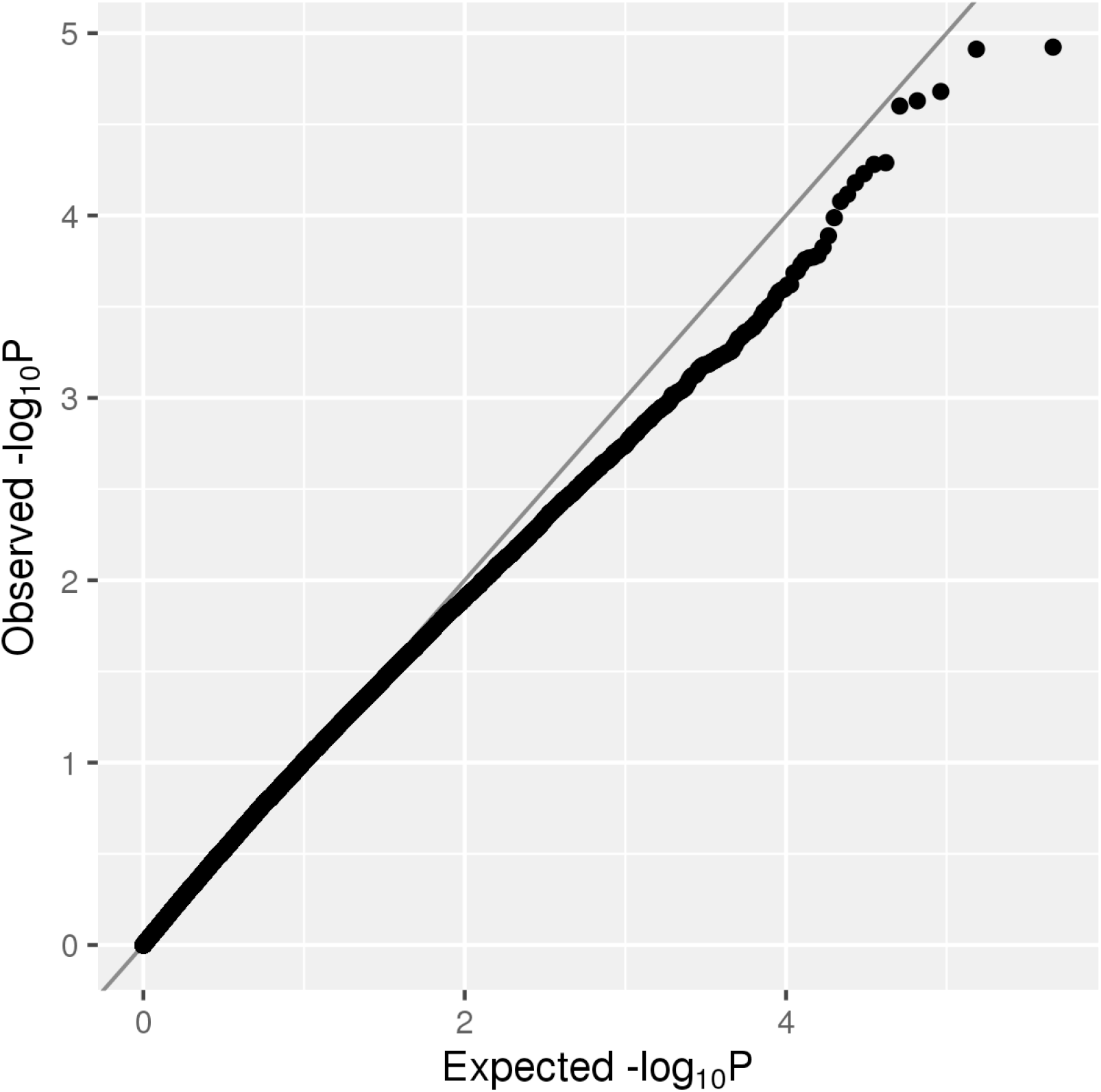
QQ plot of spatial clustering association results based on rare (MAF<=0.01) variants in the family-based discovery dataset (NIMH and NIA cohorts).

Finally, we also performed gene-based burden testing on rare variants in known AD genes (i.e. *APP, PSEN1, PSEN2* as well as those recently highlighted as genome-wide significant loci in GWAS (Jansen *et al*.^*3*^ and Kunkle *et al*.^5^). This revealed two nominally significant association signals in *ZCWPW1* (*P* = 0.028) and *PICALM* (*P* = 0.03) and two suggestive association signals in *ALPK2* (*P* = 0.053) and *MS4A6A* (*P* = 0.084), upon meta-analysis (Supplementary Table 8).

For comparison, we also plotted spatial-clustering-based association results without MAF restriction in the discovery cohorts, i.e. for both rare and common variants and, as expected, the top-associated region in these analyses maps to the *APOE* locus on chromosome 19q13.32 (Supplementary Fig. 5 and 6, and Supplementary Table 9).

**Fig. 6.**
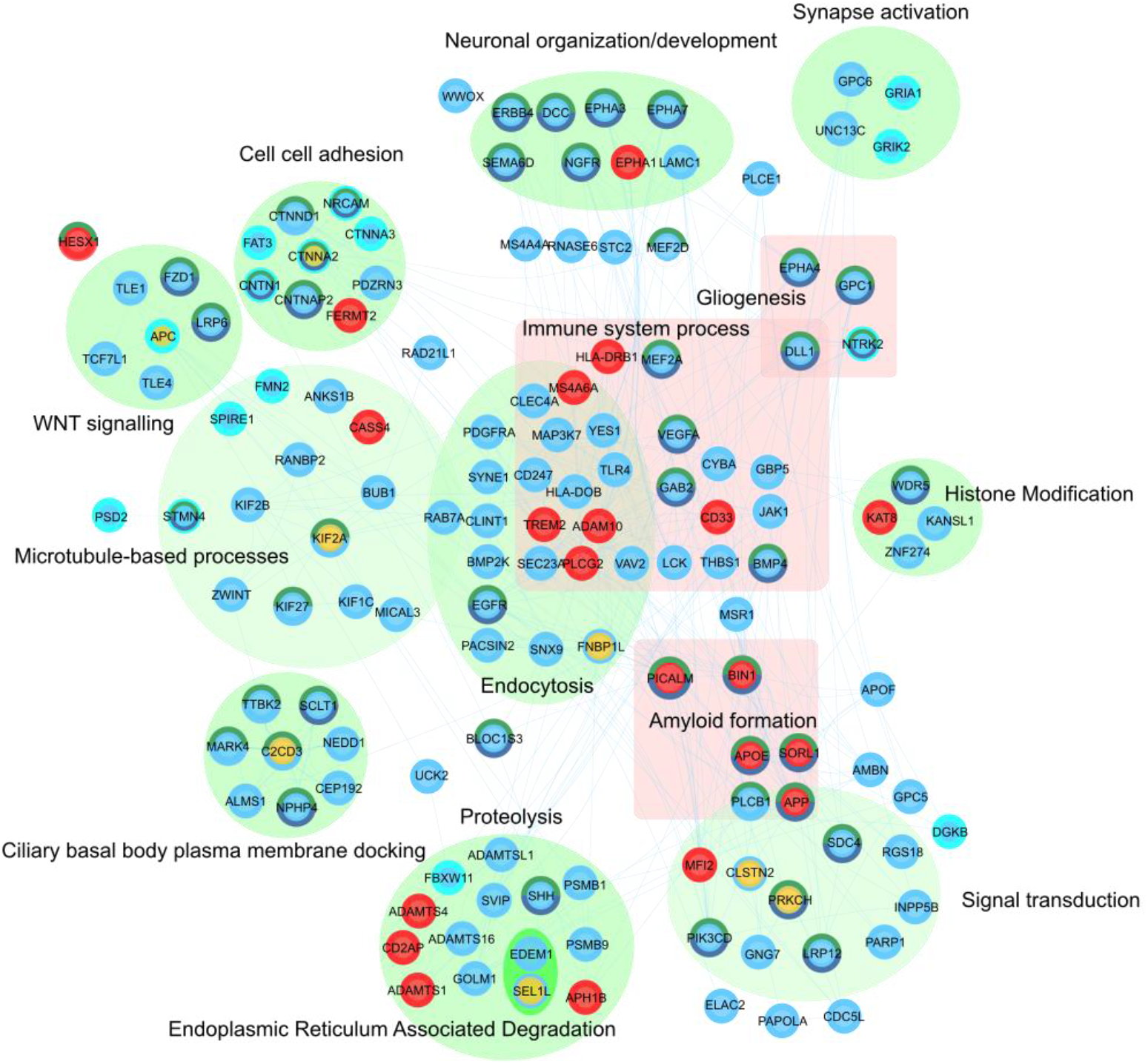
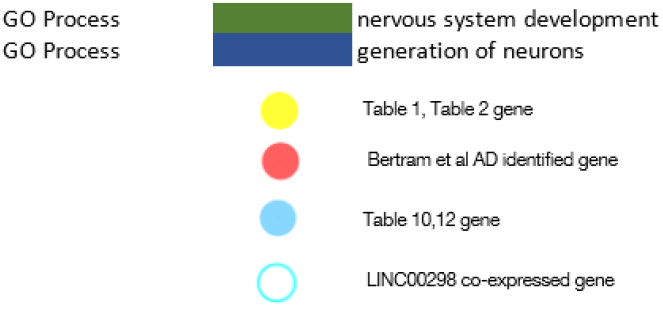
Network of direct interactions between highly ranked SNV and regional genes and known AD- associated genes. Direct protein-protein relationships (blue links) between reference AD genes (red), Table 1 and 2 (yellow), Supplementary Table 11 and 14 (blue) protein-coding genes. *LINC00298* co- regulated expression of directly interacting genes is highlighted (turquoise border). Proteins that are in direct interaction with genes from Table 1 and 2 have been grouped where possible according to shared GO biological processes (green ellipse). Proteins that may not be directly interacting but are found commonly enriched in immune-related processes are grouped (pink square). Proteins with dark green-colored borders are enriched in GO:BP *nervous system development* while a navy blue border is enriched for *generation of neurons*. Gene-gene relationships are listed in Supplementary Table 22. The network can be interactively explored via the NDEX project website (https://tinyurl.com/y6p9xjlw).

Taken together, our WGS-based association results revealed 13 novel potential AD loci (4 from single-variant, 9 from spatial-clustering analyses) with consistent rare-variant signals in both discovery and replication cohorts. Importantly, none of the identified loci have been previously highlighted in any common variant or WES/exome-chip association study in the field, emphasizing the added resolution and power afforded by genome-wide sequencing performed outside coding regions. Notwithstanding, some of the loci highlighted here may reflect spurious associations due to type-I error; thus, any future consideration of our results should await further validation in independent samples.

### *In silico* functional implications of the single-variant association findings

The leading SNV associations from the discovery (Table 1a) and the meta-analysis (Table 1b) include rs74065194, which is upstream of *SEL1L*, located within a transcription factor binding site cluster. The SNV rs192471919 is situated within the intron of *FNBP1L* and open chromatin specific to the brain cingulate gyrus, liver cells and monocytes. Three SNVs are intronic to *LINC00298* (rs147918541), a long non-coding RNA gene mostly expressed in brain, and *C15orf41* (rs147002962; rs141228575) which is mostly expressed in heart. The four variants assigned to *STK31*, which almost reach our *P*-value threshold, show significantly higher expression in the temporal cortex of AD patient samples when compared to controls (*P*_adj._ = 1.1×10-5). Of these SNVs, rs112941445 is the most likely to be the causal variant given that it has the most epigenetic support (Table 1a).

The most highly significant SNV-associated meta-analysis gene was *LINC00298*. This long intergenic non-coding RNA (lincRNA) has no known function^20^, but exhibits CNS-specific expression, with a 50-fold and 24-fold enrichment in the nervous system and brain samples in FANTOM 5 CAT (*P* = 2.9×10-23 and 4.6×10-21 respectively)^21^, including iPSC-derived neurons. Orthologous transcripts to *LINC00298* can be only be found in primates (∼45% exonic identity in *Pan troglodytes, Chlorocebus sabaeus, Papio anubis)*, and with brain-specific expression^22^. A resource for experimental characterization of lincRNA (lncBase v2 database), reveals that *LINC00298* contains an experimentally supported microRNA binding site for miR- 7, discovered by brain high-throughput sequencing of RNA isolated by crosslinking immunoprecipitation (HITS-CLIP) experiments^23^. miR-7 is expressed highly in the brain and has been implicated in numerous mechanisms in neurodevelopment, healthy brain function as well as in brain diseases, including AD, neuroinflammation, Lewy body dementia, psychiatric disorders, and Parkinson’s disease^24^. Of the 73 rare-variant-associated genes co-expressed with *LINC00298* in our study, *17* are included in protein-protein interactions with our newly AD-associated genes, including *APC* (corr 0.449) and *CTNNA2* (corr 0.381) (Fig. 6), and also the known AD and frontal lobe dementia-associated gene encoding tau protein, *MAPT* (corr 0.379). Functional enrichment for *LINC00298*-correlated expression of genes found in our study results in one significant enrichment for the *HIPPO signaling pathway* (*P*_adj._ = 2.2×10-7; Supplementary Table 10) and weaker correction-adjusted significance for GO processes *synapse organization, spindle formation, cell-cell adhesion*, and *neuron projection morphogenesis*.

Functional enrichment of the genes associated with the highest-ranked 1000 SNVs from the meta- analysis (Supplementary Table 11) identified 151 processes and pathways after correcting for multiple testing. The most highly enriched terms included *flavonoid glucuronidation* (*P*_adj._ = 1.09×10-7) (involved in removal of xenobiotics), and many neuroplastic/developmental-associated processes including *synapse organization* (*P*_adj._ = 1.32×10-7), *axon guidance* (*P*_adj._ = 6.51×10-6), *development* and *elongation*, and also *cell adhesion* (*P*_adj._ = 0.001; Supplementary Table 12). Only two pathways were significantly co-enriched with the GO/pathway gene set enrichment for genes associated with common variants reported in the GWAS by Jansen *et al*.^*3*^: ce*ll adhesion molecules* and *herpes simplex infection* (Supplementary Table 13). In contrast to the broad diversity of functions, such as immune-related and amyloid processing, found to be enriched by genes annotated in the GWAS by Jansen *et al*.^*3*^, 10 of the 21 top-level functions showing enrichment in our rare-variant analysis had roles related to the maintenance and development of neurons, cardiac tissue and synapses, and neuroplasticity-related terms including *synaptogenesis, activity and synaptic integrity, neurogenesis, sensory organ development, cardiac development, tissue morphogenesis*, and *limb development*. None of the enriched pathways, here, exhibited immune-related roles.

### *In silico* functional implications of the spatial-clustering association findings

Four of the nine leading regions associated with AD are significantly enriched for regulatory annotation (Table 2). The *CLSTN2* and *PRKCH* regions are respectively enriched for enhancers and promoters across a number of cell types, while the *LHX9* and *NALCN* loci significantly overlap with transcription factor binding sites. *NALCN* additionally is enriched for active CTCF binding sites. Unlike the SNVs, these nine regions mostly cover intronic and exonic locations. The four genes *APC, CTNNA2, KIF2A*, and *NALCN* are all primarily expressed in brain tissue while *PRKCH* expression is significantly reduced in the temporal cortex of AD patients (*P*_adj._ = 0.0001).

Functional enrichment of genes associated with the highest-ranked 1000 spatial clustering-based results (Supplementary Table 14) revealed 127 significantly enriched pathways after correcting for multiple testing. The most highly enriched terms included *neuron projection guidance* (*P*_adj._ = 1.6×10-5), *kidney development* (*P*_adj._ = 2.32×10-5), *cell-cell adhesio*n (*P*_adj._ = 6.53×10-5), *negative chemotaxis* (*P*_adj._ = 2.17×10- 4), *brain development* (*P*_adj._ = 4.23×10-4) and *synapse organization* (*P*_adj._ = 7.02×10-4). 7 of the 20 most enriched terms were related to development, and 81 of the total significantly 127 enriched terms related to development or neuroplasticity (Supplementary Table 15). Meanwhile, no process out of 422 was significantly enriched in common with the Jansen *et al*. study^3^. *Protein localization to membrane* (*P*_adj._ *=* 0.0126, Jansen, *P*_adj._ = 0.0631, regional geneset; Supplementary Table 16) was the closest to reaching significance.

### Common functional themes between single-variant and regional analysis

A total of 90 genes were found in common between the most highly ranked 1000 single-variant and regional findings. These include three highlighted genes *LINC00298, SEL1L*, and *STK31* and genes that rank highly in both gene lists: *ROBO1, PRDM9, LINC02439*, and *TMEM132C*. 152 processes and pathways reached significance for the co-enrichment of regional- and SNV-associated genes. The top 5 terms enriched were *positive regulation of nervous system development* (*P*_adj._ = 0.0025, SNV; *P*_adj._ = 0.000079, regional), *heart development* (*P*_adj._ = 0.0015, SNV; *P*_adj._ = 0.0039, regional), *sensory organ development* (*P*_adj._ = 0.0005, SNV; *P*_adj._ = 0.01, regional), *trans-synaptic signaling* (*P*_adj._ = 0.0015, SNV; *P*_adj._ = 0.0031 regional) and *tissue morphogenesis* (*P*_adj._ = 0.002, SNV; *P*_adj._ = 7×10-5 regional). Of the 19 significantly co-enriched terms, 10 were related to development or neuroplasticity, the remainder addressed maintenance and cellular activity-related functions such as *cell-cell adhesion, negative chemotaxis, signaling by receptor tyrosine kinases* and *organelle localization* (Supplementary Table 17; Supplementary Fig. 7).

**Fig. 7.**
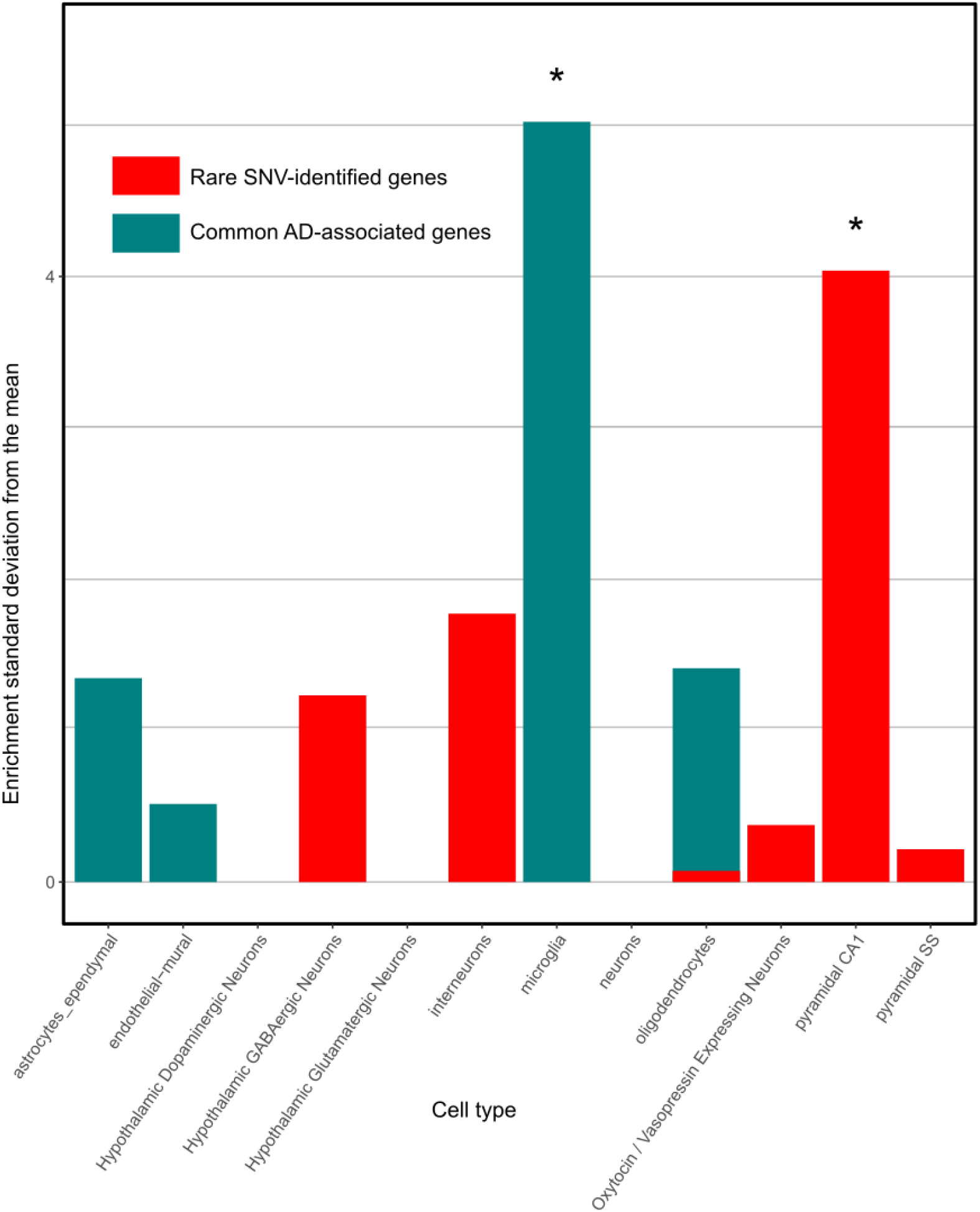
Cell-specific enrichment results from the EWCE tool. We compared genes identified in our rare-variant analysis to common variants published in AD^26^ and which cell type each is significantly enriched in. Zero represents the mean expression in each cell based on 10,000 permutations of gene lists of the same size. The data for this figure can be found in Supplementary Table 18.

To investigate the impact of selecting only variants within transcribed gene boundaries on the functional enrichment results, we restricted our analysis to only those SNVs and regions occurring within a gene transcript, i.e. intronic or exonic variants. The most highly enriched categories (*P*_adj._ <= 0.035) included *organelle localization, cell-cell adhesion, cell morphogenesis in neuron differentiation, synapse organization, modulation of chemical transmission*, and *protein localization to the centrosome* (Supplementary Fig. 8).

### Identification of cell-specific signatures

To assess whether our prioritized variants show an association with single-cell-restricted states, we applied an Expression Weighted Cell Type Enrichment (EWCE) test^25^ to genes from our prioritized SNV and regional analysis results. EWCE is used to predict the primary cell origins of a disease. Using single-cell mouse data, primarily from the hippocampus and hypothalamus, we discovered an enriched signal of our SNVs in pyramidal CA1 neurons (Supplementary Table 18). In contrast, common loci associated with AD^26^ have been significantly enriched in microglia (Fig. 7).

### Network generation of shared functions and relationships with known AD-associated genes and processes

Using known protein-protein interactions as a guide, a network of interactions was constructed between a total of 1,274 interacting proteins which include known AD-associated genes26, our single-variant, and regional-associated genes. Of the 14 leading genes we pinpointed in this study, 8 (protein-coding) were linked directly by protein-protein interaction to additional AD-associated genes discovered within this study or to 21 known AD-associated genes in a subnetwork (Fig. 6). Highlighted genes that interact with known AD genes include *FNBP1L*, which directly interact with the validated GWAS AD genes, *PICALM* and *BIN1*, as well as *KIF2A*, which directly interacts with AD gene *HLA-DRB1*. 17 genes in the subnetwork also co-express with the highlighted gene, *LINC00298*.

Functional enrichment of the subnetwork of directly interacting proteins revealed 196 enriched GO process/KEGG pathway terms (Supplementary Table 19). The 3 highest ranked GO processes (*nervous system development*, 236 genes, FDR 1.32×10-9; *neurogenesis* 168 genes, FDR 4.74×10-7; *developmental process*, 460 genes, FDR 3.74×10-7) reflected neuroplasticity/developmental processes, of 90 processes enriched for development, differentiation, or biogenesis. *Neurogenesis*, a GO process term that annotates 1,519 genes, was co-enriched with *PRKCH, LHX9* and *CTNNA2* from our pinpointed genes, and *SORL1, PICALM, CNTNAP2* and *APOE*, and *BIN1* from our reference list of known AD genes (*PICALM* is a known AD gene also discovered in our top 1000 regional analysis-associated genes). Co-expression analysis using pathway co-activation mapping (PCXN.org^27^) revealed that *nervous system development* and several of the associated enriched GO terms show significant correlated gene expression activity, even when there was low gene overlap between enriched term genesets (Supplementary Table 20).

## Discussion

Based on WGS of 2,247 subjects from 605 multiplex AD families and a case-control cohort of >1,650 individuals, we have identified 13 rare-variant signals (4 from single-variant, 9 from spatial-clustering analyses) exhibiting association with AD across the discovery (families) and replication (case-control) cohorts. Our work represents one of the first and, to the best of our knowledge, the currently largest, systematic WGS-based genetics study in the AD field. In AD, we are only aware of two published WGS-based studies^28,29^ both utilizing different analyses paradigms and much smaller sample sizes. Of note, data from the latter of these WGS projects were utilized in the current study for purposes of independent replication.

The top signals emerging from our single variant-associated analyses were associated with the genes: *FNBP1L* and *SEL1L (*and *STK31*), while the secondary analysis pointed to *LINC00298* and *C15orf41*. All genes directly overlapped with the single variant associations except for *SEL1L*, which encodes the suppressor/enhancer of lin-12-like (Sel1L) adaptor protein for an E3 ligase involved in endoplasmic reticulum-associated degradation (ERAD) for protein quality control. Interestingly, ERAD has been reported to regulate the generation of amyloid-beta by gamma secretase^30^. Deficiency of *SEL1L* has also been show to activate ER stress and promote cell death^31^. Additionally, an SNV in intron 3 of *SEL1L* has previously been reported to confer susceptibility to AD^32^.

The *FNBP1L* gene, which encodes the formin-binding protein 1-like protein, has been associated with adult^33^ and childhood intelligence^34^. *FNBP1L* has also been reported to be essential for autophagy of intracellular pathogens, such as *Salmonella Typhimurium*, which serves to curb intracellular growth^35^. This is particularly interesting given the emerging evidence for the role of microbes in driving AD neuropathology^36^. *FNBP1L*, also known as *TOCA-1*, is implicated in neurite elongation and axonal branching^37^. Thus, *FNBPL1* may play a role in neuroplasticity-related AD pathology.

The *STK31* gene encodes the cell cycle kinase, serine/threonine kinase 31, which is known to promote *PDCD5*-mediated apoptosis in p53-dependent human colon cancer cells^38^. It is tempting to speculate as to whether this kinase might also affect phosphorylation of tau and neurofibrillary tangle formation in AD. However, we note that variants in this gene technically did not fulfill the significant thresholds and are highlighted here as additional results.

*LINC00298* is a long intergenic non-coding RNA and does not code for a protein. Its functional role is not known. It contains a target for brain-expressed non-coding miRNA mir-7, which has been associated with AD^39^. *LINC00298* can be more broadly functionally characterized by where and when it is expressed and the genes with which its expression is correlated. *LINC00298* is co-expressed with 33 SNV-associated, and 40 regional-associated genes (lnchub^40^). *LINC00298’s* co-expressed genes appear to be enriched for developmentally-associated processes: Its bias for expression in the brain, its association with HIPPO pathway, which has a role in development, co-expression with genes involved in neuronal differentiation and expression in iPSC neuronal stem cells suggest that one of its roles may be in regulation involved in neuronal plasticity. *C15orf41* encodes the codanin 1-Interacting nuclease gene (*CDIN1*), which is highly expressed in the heart, with much lower expression in the brain. *CDIN1* is associated with erythrocyte differentiation and has genetic associations with congenital dyserythropoietic anemia type I^41^.

Spatial clustering-based analyses highlighted a total of four independent genomic regions (Table 2a). One of these regions was in the gene encoding the protein kinase C receptor beta subunit (*PRKCH*). Interestingly, we have previously reported three highly penetrant rare mutations in another protein kinase C subunit alpha (*PRKCA*) that segregates with AD in five families. All three AD-linked *PRKCA* mutations displayed increased catalytic activity (by live imaging) versus wild-type *PRKCA*, and potentiated the ability of amyloid beta to suppress synaptic activity in hippocampal slices^42^. It will be interesting to determine whether mutations in *PRKCH* have similar aberrant effects on receptor activity.

The three other genes implicated in the spatial clustering-based analyses included *C2CD3*, which encodes the C2 domain containing 3 centriole elongation regulator that is expressed at relatively high levels in the brain. Mutations in human *C2CD3* cause skeletal dysplasia, caused by defective assembly of the primary cilium, a microtubule-based cellular organelle involved in developmental signalling^43^. *KIF2A* encodes the kinesin family member 2A, which is required for normal mitotic spindle activity and normal brain development, most likely via its ATP dependent MT-depolymerase activity^44^. Like *C2CD3, KIF2A* has also been implicated to affect ciliogenesis, relating to its role in the cell cycle. *KIF2A*-related cortical development defects have been attributed to decoupling between ciliogenesis and cell cycle^44^. A *KIF2A* His321Asp missense mutation was identified in a subject with defective cortical development owing to impairment of KIF2A microtubule depolymerase activity^45^. Several members of the kinesin family are overexpressed in the brains of AD patients^46^, and *KIF2A* expression is specifically upregulated in axons, spinal neurons, and oligodendrocytes adjacent to spinal cord injuries^47^ Finally, *APC* encodes the Adenomatosis Polyposis Coli Regulator of WNT Signaling Pathway (as a negative regulator) and serves as a major tumor suppressor. The WNT signaling pathway plays an important role in the development of the central nervous system, including axonal pathfinding and synaptic plasticity, and has been linked to AD pathogenesis^48^. Aβ neurotoxicity in AD has been reported to downregulate WNT signaling^49^, and WNT signaling, in turn, has been shown to regulate β-secretase cleavage of APP^50^. Collectively, these findings indicate that inhibition of WNT signaling may play a role in the generation and neurotoxicity of Aβ. Thus, *APC* may influence AD neuropathogenesis via regulation of the WNT signaling pathway.

In addition to these four loci, an additional five candidate regions were identified in the secondary analyses, based on the top meta-analysis results (*P*<5×10-5; Table 2b). These included *LHX9, NALCN, CTNNA2, SYTL3*, and *CLSTN2. LHX9* is a LIM homeobox gene family member and is involved in the development of the forebrain^51^. This gene has also exhibited genetic association with “self-reported educational attainment”^52^. *NALCN* encodes a voltage-gated sodium and calcium channel that is expressed in neurons. Interestingly, the calcium-sensing receptor, CaSR, which has been reported to regulate *NALCN*, has been previously implicated as an important signaling molecule in AD^53^. *CTNNA2* encodes the neural version of α-catenin (αN-catenin), a mechano-sensing protein that links cadherins with the cytoskeleton; as such, they are required for proper neuronal migration and neuritic outgrowth^54^. *SYTL3* encodes the Rab effector protein, synaptotagmin-like 3, which plays a role in vesicle trafficking^55^, and has been genetically associated with lipoprotein (a) levels^56^. *CLSTN2* encodes Calsyntenin 2, which modulates calcium-mediated postsynaptic signaling in the brain. Absence of *CLSTN2* impairs synaptic complexes in mice^57^, and has been associated with episodic memory function in human subjects^58^.

Pathway analyses based on our highlighted rare-variant-associated genes, emphasize functional roles in neuroplasticity, synaptic function and integrity, axonal maintenance, neuronal development, and heart tissue development. In contrast, genes identified through common-variant associations by GWAS have been more involved with pathways linked to immune-system response, lipid metabolism, and Aβ deposition. This stark difference in enrichment profiles may represent an essential contribution of rare variants to the development of AD based more on neuronal and synaptic function. This finding is further substantiated by examining our SNV-associated genes and published common AD-associated genes for cell-specific biases in expression. We found that hippocampal CA1 neurons were significantly enriched for our rare signature whereas common genes from AD GWAS have primarily highlighted microglia as the likely primary cell type of effect (Fig. 7).

Using whole-genome sequencing, we have performed a whole-genome global screen to search for association of rare variants with Alzheimer’s disease. It is noteworthy that our most significantly SNV-associated gene, *LINC00298*, is non-coding and of unknown function. Furthermore, all nine regions of the genome we have identified to be associated with AD risk, overlap with regulatory annotations, of which four are significantly enriched. Thus, our study emphasizes the importance of focusing on the non-coding part of the genome for a better understanding of the genetic and functional basis of Alzheimer’s disease.

The methodologies applied and the results obtained are not without limitations. First and foremost, we note that the size (n∼2,300) of the discovery sample is relatively small compared to common-variant GWAS in the field. This is due to the limited availability of samples (i.e. multiplex AD families) and funds (i.e. costs for generating WGS data are still 1-2 orders of magnitude higher than for common-variant GWAS which rely on microarray-based genotype calls). This increases both the type I (i.e. the chance of false positives) and type II (i.e. chance of false negatives) error rates of our study. We tried to alleviate this limitation by utilizing validation data from an independent case-control WGS dataset (NIA ADSP), but all of the main findings highlighted here should be considered preliminary until validated in additional datasets.

Second, most variants highlighted to be associated with AD risk in our analyses are located in non-coding regions of the genome. While this is to be expected given the proportions of coding (∼2%) vs. non-coding (∼98%) sequence variation in humans, it aggravates efforts to validate and functionally annotate our top findings. However, efforts like ENCODE (http://www.encodeproject.org), the NIH Epigenomics Roadmap Consortium (http://www.roadmapepigenomics.org/), or the International Human Epigenome Consortium (http://ihec-epigenomes.org/) continue to provide compelling evidence that an increasing fraction of disease-associated variation maps to the regions between genes, providing a strong argument for using whole-genome in addition to whole-exome approaches to capture the full rare-variant architecture underlying AD.

Finally, unlike genetic association analyses in case-control settings, our family-based approach is robust against common genetic confounders due to population substructure. However, given the fact that more than 80% of our discovery family-based sample were individuals of European ancestry, we limited our replication sample to individuals of the same ancestry. This comes at the price of reduced statistical power which we addressed by adjusting the discovery and meta-analysis significance thresholds. As a result, our top findings show *P*-values ranging between ∼0.01 and ∼8×10-7, which is still almost two orders of magnitude above a recommended threshold (*P*<1×10-8) for rare-variant-based studies in European-based samples^59^. Eventually, only the generation and analysis of additional datasets investigating these and other rare variants in relation to AD susceptibility will allow us to distinguish true from false-positive findings.

In summary, here, we describe the first WGS-based rare-variant association study in AD, and highlight several novel variants and regions found to be associated with disease risk. Subsequent functional annotation assessments imply several molecular pathways to be relevant in AD based on rare variant analysis, e.g. neuronal development and synaptic integrity. This contrasts with innate immune and lipid pathways previously implicated by network analyses of AD GWAS based on common variants. Together with the results of common-variant AD risk GWAS, our study highlights several novel promising routes of AD research and provides new potential targets for therapeutic interventions aimed at the early treatment or prevention of AD.

## Methods

### Sample descriptions

The discovery cohort was composed of two WGS familial cohorts with 1,393 (NIMH; AD: n=966) and 854 (NIA ADSP families; AD: n=543) individuals. A subject was considered to be affected if he/she was included in these categories: “definite AD”, “probable AD” or “possible AD”. Unaffected subjects were taken from one of the following categories: no dementia (667 subjects), suspected dementia (46 subjects) or non-AD dementia (10 subjects). It is important to note that NIA ADSP families by design did not include individuals with two APOE-ε4 alleles. Since our discovery cohort consisted of mostly individuals of European ancestry, we used a matching subset (non-Hispanic whites [NHW]) from the replication cohort (NIA ADSP unrelated, n=1669). A total of 564 individuals (AD: n=307) were obtained with RNA-Seq data in the temporal cortex from Mayo Clinic Alzheimer’s Disease Genetics Studies (MCADGS^60^). All datasets are described in Supplementary Table 1.

### Whole-genome sequencing methods

Plated DNA was obtained from the Rutgers Cell Repository and sent to Illumina Inc (San Diego, CA, USA) and used to create short-insert paired-end libraries. Paired-end libraries are manually generated from 500ng–1ug of gDNA using the Illumina TruSeq DNA Sample Preparation Kit. Samples are fragmented and libraries were size selected targeting 300 bp inserts and sequenced using the HiSeq 2000 System. Illumina-provided BAM files were re-aligned to the human reference genome (GRh38) with bwa-mem^61^ (v0.7.7, default parameters). Reads were marked for duplication using samtools^62^ (v0.1.19). Germline variants were jointly called for each family using FreeBayes^63^ (v0.9.9.2-18) and GATK^64^ (v3.0) best practices method (https://software.broadinstitute.org/gatk/best-practices/) as part of the bcbio-nextgen workflow (https://github.com/chapmanb/bcbio-nextgen) before being squared-off with bcbio.recall (https://github.com/chapmanb/bcbio.variation.recall) across the whole cohort to distinguish reference calls from no variant calls. Library and read quality was assessed using FastQC (v0.10.1; http://www.bioinformatics.babraham.ac.uk/projects/fastqc/) and Qualimap^65^ (v0.7.1).

### Quality control of WGS-derived variant calls

We first performed individual-based quality control. Based on genotyping rate and inbreeding coefficient we removed three outliers in the NIMH dataset. Further 12 duplicates and 24 individuals with wrong family assignments, as per estimated identity by descent (IBD) sharing, were removed as well (Supplementary Table 20). 1,393 clean individuals from NIMH were combined with 854 individuals from NIA and analysis was performed only on variants present in both datasets. This was done to ensure a consistent discovery dataset for region-based rare-variant analysis. Next, family-based discovery datasets were filtered for monomorphic variants, singletons, variants with a missingness rate higher than 5%, Mendelian errors, and variants which had a Hardy-Weinberg equilibrium *P*<1×10-8. Only variants which had a filter “PASS” in the vcf file were included in the analysis.

In the case-control replication datasets, variant-based filtering was performed as in family-based datasets, i.e. monomorphic variants, singletons, variants with a missingness rate higher than 5% and variants that had a Hardy-Weinberg equilibrium *P*<1×10-8 were excluded. Only variants that had a filter “PASS” in the vcf file were included in the analysis. We kept only unrelated individuals of European ancestry, in order to closely match our discovery dataset population. Principal components were calculated based on rare variants using the Jaccard index^66^. Outliers based on principal components were excluded.

### External minor allele frequency reference dataset (gnomAD)

We have downloaded v3 of the Genome Aggregation database (gnomAD)^12^, which included 71,702 whole genomes (32,399 non-Finnish European). For minor allele frequency we used the AF NFE field, which corresponds to allele frequency in the non-Finnish European population. Variants were considered rare, if AF NFE was less than 1% or more than 99%.

### Single-variant association analyses

In the family-based discovery datasets we used the FBAT Toolkit^14^ to perform association analysis on variants seen in at least one informative family in combined NIMH/NIA dataset. We used an offset of 0.15 which approximately corresponds to the population prevalence of disease. In the case-control replication datasets we performed a logistic regression (with option “firth-fallback”) for case/control status as implemented in PLINK 2^67^. We included sex, age, sequencing center and 5 Jaccard principal components^66^ with standardized variance as covariates. We next performed a fixed-effects meta-analysis of 2 datasets. The meta-analysis was performed with the METAL toolkit^68^ with a sample-size-based weighting scheme. Quantile-quantile plots were drawn in R for all results and for variants with at least ten informative families.

### Spatial-clustering/region-based association analyses

In the family-based discovery dataset, we systematically grouped the whole-genome sequencing data into non-overlapping regions using a spatial-clustering approach^19^. Briefly, regions include variants which are in close proximity to each other. We included only variants, seen in at least two families. After partitioning the chromosomes into non-overlapping windows. FBAT-RV^69^, which is a multimarker test with minor allele frequency (MAF) weighting, was used to test identified non-overlapping regions in the combined family-based dataset. First, only rare variants were included in the analysis. Next, we performed a second run including all variants.

In the case-control replication datasets, joint variant testing was performed on rare variants using the burden test as implemented in the SKAT package^70^. We next used SKAT-RC^71^ to incorporate all variants with no MAF threshold. We used the same set of covariates as in the single-variant analysis. For consistency, we tested the same non-overlapping regions, which were identified in the combined NIMH/NIA dataset. This allowed us to perform a meta-analysis of the identified regions, using Fisher’s combined probability test.

### Variant and regional association with genes

Disease-associated variants are often assigned to genes by their close proximity, where only genes overlapping or closely flanking the reported SNVs are considered. The overlap-only strategy excludes other potentially causal genes within the associated haplotype. However, expanding gene association to include non-overlapping SNVs or regions is complicated by the current diversity and inconsistency of annotation for non-coding regions of the genome. As regulatory regions proximal and distal to a gene are becoming extensively annotated^72^, we have leveraged the functional significance of sets of cis-regulatory regions of the vertebrate genome. We applied The Genomic Regions Enrichment of Annotations Tool (GREAT) to leverage functional cis-regulatory regions identified by localized measurements of DNA binding events across the genome^73^. Applied to non-gene overlap regional and SNV loci; GREAT associated additional genes to both SNVs and regions.

### Differential gene expression

A mixed effect linear regression was performed on the RNA-Seq output with Bioconductor (v3.7) using CQN^74^ and limma^75^ adjusting for clinical and technical variations. A multiple testing correction was applied.

### Annotation and geneset enrichment

Prioritized variants and regions were annotated for relationships to eQTLs (GTEX^76^), CpG islands, DNase hypersensitivity, RNA gene locations and RNA binding sites (UCSC^77^), enhancers, promoters, transcription start sites, transcription factor binding sites and other regulatory features (Ensembl^78^; FANTOM5^79^), histone marks and GC-content (GWAVA^80^), 3D genomic interactions and open chromatin (3DSNP^81^), cell-specific enhancers (INFERNO^82^) and the Illumina bodyMap2 transcriptome (GSE30611).

### Regulatory enrichment within spatial-clustering/region-based association

To test whether the top regions of interest were overpopulated with regulatory annotations, we computed 103 random permutations per region, across the genome of the same length to count the number of overlapping annotations. These regions were restricted to regions with similar numbers of genes. A fisher’s exact test was used to compare annotations within the top leading regions against these permuted regions. Multiple testing correction was applied for every region x annotation that was tested.

### Cell-specific enrichments

We performed Expression Weighted Cell Type Enrichment with EWCE^25^ using mouse single-cell transcriptomic data from the cortex and hippocampus^83^. EWCE aims to identify the cellular origins of a disorder by examining where a disease-associated gene list is primarily expressed and testing this against a distribution obtained from 10,000 permutations of random lists. We selected four gene lists to be tested: the leading SNV/region-associated genes from Table 1 and 2 (n=5), SNV-associated genes (*P*_meta_ < 0.01; n=185), region-associated genes (*P*_meta_ < 0.0005; n=55), and published common-variant AD-associated genes^26^ (n=32). 78% of these genes had a mouse homolog which were then used in the analysis.

### Functional enrichment analysis for associated genes

Functional enrichment for the SNV- and regional-associated genes or for genes found to be co-expressed with *LINC00298*, was performed via the Metascape server^84^ which applies the hypergeometric test^85^ and Benjamini-Hochberg *P*-value correction algorithm^86^ to identify terms (all GO ontologies, Reactome and KEGG pathways) that contain a statistically greater number of genes in common with an input list than expected by chance. Enriched terms were filtered at an FDR <= 0.1.

### Network relationships with known AD genes

First, we set out to understand novel but direct relationships between genes associated with our identified variants and regions and already published Alzheimer’s-associated genes. These known Alzheimer’s genes were selected from a recently published review^26^ and include genes which cause familial forms of the disease (e.g. *APP, PSEN1* and *PSEN2*) as well as genes which have the highest association in GWA studies^3,5,9,87,88^. We used the StringDB protein-protein interaction resource^89^ using only identified protein-protein interactions. Using this background that agglomerates protein-protein interaction datasets, we identified direct (curated AD genes directly interacting with our associated genes) associations in a global network which contained 22 known AD genes, 73 regional-associated genes and 59 SNV-associated genes (Supplementary Table 22). This network was reviewed for direct interactions between known AD genes and SNV/regional associated genes. Genes related to each other in this manner were then visualized using Cytoscape^90^. Genes in this network co-expressed with *LINC00298* were highlighted when correlated in expression as defined according to pre-calculated correlations available at the lncHUBhub server (https://amp.pharm.mssm.edu/lnchub/; Supplementary Table 23). The server provides gene-lncRNA correlation computed from 11,284 TCGA normalized samples processed by recount2^40^, gene counts are quantile normalized and the Pearson correlation is computed.

Functional enrichment within this network was performed using the remote StringDB server linked to Cystoscape “String App Enrichment function”^91^, producing enrichments using the hypergeometric test, with *P*-values corrected for multiple testing using the method of Benjamini and Hochberg in known molecular pathways and GO terms as described in Frenceschini *et al*.^92^. Enriched GO/pathway terms were considered at an FDR <=0.05. Genes from our study and known Alzheimer’s genes coding for proteins directly interacting with proteins identified by genes from Table 1 and Table 2 were examined for common enrichment and grouped around the genes we highlighted in these tables into functional clusters where possible. Genes from our study or known AD genes which show protein-protein interaction links with Table 1- and Table 2-identified genes were grouped most closely in the common annotation clusters. The top GO enrichment classes (*nervous system development* and *generation of neurons*) were annotated to nodes using the *String enrichment color palette* function to produce highlighted node borders. Immune-related functions which showed enrichment for currently known AD-related genes were used to group both known AD genes and regional-associated and SNV-associated into annotation clusters.

## Supporting information

Main and supplemental tables

## Data Availability

Data available upon request. Summary statistics will be made available after peer-review.

## Acknowledgements

The authors would like to thank the staff from the National Institute of Mental Health (NIMH) Divisions of Clinical and Treatment Research (DCTR) and Epidemiology and Services Research (DESR), including David Shore, MD, Mary Farmer, MD, MPH, Debra Wynne, MSW, Steven 0. Moldin, PhD, Darrell G. Kirch, MD (1989-1994), Nancy E. Maestri, PhD (1992-1994), William Huber (1989-1995), Pamela Wexler (1995-), and Darrel A. Regier, MD, MPH. They would also like to thank the study staff at all three sites and the data management staff at SRA Technologies, Inc., particularly Cheryl McDonnell, PhD, for the care and attention that they paid to all aspects of the study. The authors are also extremely grateful to the families whose participation made this work possible. We would like to thank Dr. Ioannis Vlachos and Leinal Sejour, Beth Israel Hospital Non-coding RNA precision diagnostics and therapeutics core of the Harvard Medical School Initiative for RNA Medicine, Beth Israel Deaconess Medical Center, for their help in interpretation of non-coding genome and ncRNA genes.

This study was supported by the Cure Alzheimer’s Fund, and the following federal grants: U24AG026395 (NIA-LOAD Family Study); U24AG021886 (National Cell Repository for Alzheimer’s Disease); P50AG08702 (Boston University and Columbia University); P30AG028377 (Duke University); P30AG010133 (Indiana University); PO1 AG05138 (Massachusetts General Hospital; Mayo Clinic, Rochester; Mayo Clinic, Jacksonville; and Mount Sinai School of Medicine); and P30AG010124 (Northwestern University Medical School; Oregon Health and Science University; Rush University Medical Center; University of Alabama at Birmingham; David Geffen School of Medicine, University of California, Los Angeles; University of Kentucky, Lexington; University of Pennsylvania; University of Pittsburgh; University of Southern California; The University of Texas Southwestern Medical Center; University of Washington; and Washington University School of Medicine). This work was also supported in part by the National Institute for Health Research (NIHR) Sheffield Biomedical Research Centre (Translational Neuroscience)/NIHR Sheffield Clinical Research Facility and the Cure Alzheimer’s Fund (Alzheimer’s Disease Research Foundation) (W.A.H.). Please refer to the Supplementary Note for additional acknowledgements.

## Author Contributions

R.E.T, B.H., L.B. and D.P. designed the study. B.H., K.M., R.K., O.H. and B.C performed sequencing and quality control. D.P., B.H. and K.M analyzed the data. S.L.M., S.A. and W.A.H performed the functional analysis. O.H., W.A.H., L.B., K.M., D.P., B.H., C.L. and R.E.T. contributed ideas and insights. L.B., C.L., W.A.H. and R.E.T. supervised this work. R.E.T. and W.A.H. obtained funding. L.B., D.M. and R.E.T. wrote the original draft of the paper, and all authors edited and reviewed the manuscript.

